# Gender based disparities in Medicare physician reimbursement persist across years and specialty

**DOI:** 10.1101/2024.06.05.24308504

**Authors:** Sayuri Sekimitsu, Omar Alaa Halawa, Michael V. Boland, Nazlee Zebardast

## Abstract

**Introduction:** The gender pay gap is wide in medicine but the extent of this disparity across specialties and over time have not been elucidated. Here we evaluate differences in Medicare reimbursement between men and women physicians over time and by specialty, controlling for physician and practice characteristics.

**Methods:** The Centers for Medicare & Medicaid Services Payment Data was used to determine total reimbursements and number of services submitted by physicians practicing in the US between 2013 and 2019. Data from the American Community Survey (ACS) were used to determine average income, unemployment rates, poverty rates, income, and educational attainment levels by zip code for each physician’s practice location.

**Results:** Among the 3,831,504 physicians included in this analysis from 2013-2019, 2,712,545 (70.8%) were men and 1,118,859 (29.2%) were women. Overall, men received more in Medicare reimbursements ($58,815 ± $104,772 vs. $32,205 ± $60,556, p<0.001) and billed more services (864 ± 1,780 vs. 505 ± 1,007, p<0.001) compared to women. The median Medicare reimbursement for men decreased from 2013 to 2019 from $59,710 to $57,874, while the median Medicare reimbursement for women increased from $30,575 to $33,456. Men were reimbursed more than women across all specialties with the greatest disparity in procedure-heavy specialties. The specialties with the highest difference in median Medicare reimbursement between men and women were ophthalmology ($99,452), dermatology ($84,844), cardiology ($64,112), nephrology ($62,352), and pulmonary medicine ($47,399). In linear regression models controlling for calendar year, years of experience, total number of services, and ACS zip-code-level variables, men received a higher amount of Medicare reimbursement in all specialties, as compared to women (p<0.01 for all). The percentage of top earning men (range: 65.0%-99.5%) surpassed the proportion of men in each specialty (range: 46.1%-94.6%), except public health and preventive medicine.

**Conclusions and Relevance:** Women physicians continue to receive lower total Medicare reimbursements than men physicians, particularly in procedure-heavy specialties. Lower clinical volume and fewer procedural services among women physicians partially contribute to the disparities in reimbursement.

## Introduction

Gender differences in physician income have been documented in some specialties and settings^1–4^. Men physicians earn higher salaries than their women counterparts, even after accounting for their clinical activity and practice characteristics^4,5^. Disparities in physician reimbursement by gender are pervasive, affecting physicians in academic settings^1^, in both clinical and surgical specialties^2,5–9^, those receiving payments from Medicare (the largest single payer in the United States)^3,5^, as well as physicians practicing in other countries^10–12^. One study using data from the Center for Medicare and Medicare Services (CMS) found that gender disparities in Medicare payments to cardiologists persisted after accounting for number of charges, practice setting, patient characteristics and years of experience^5^. The study noted geographic variation in gender disparities, but women cardiologists earned less than their men counterparts in most states. While prior studies have consistently demonstrated a gender gap in physician reimbursement, even after accounting for clinical activity, patient characteristics and years of experience, few have explored longitudinal trends in these disparities or compared disparities in payments. Furthermore, while one study evaluated geographic variation in reimbursement trends^5^, the role of area-level socioeconomic conditions in gender disparities has not been explored. Finally, few studies have evaluated differences in the proportion of procedural (as opposed to office visit) services billed, and their role in disparities in total reimbursement.

In this study we aimed to evaluate gender disparities in CMS reimbursements longitudinally from 2013 to 2019 across all specialties, accounting for potential confounders including physician practice zip-code socioeconomic indicators.

## Methods

### Data Sources

This study utilized publicly available data from the CMS Physician and Other Supplier Public Use Files from 2013 to 2019^13^. The dataset provides information on services provided to Medicare Part B beneficiaries by physicians and other healthcare professionals, linked by National Provider Identifier (NPI) and Healthcare Common Procedure Coding System (HCPCS) code. Physicians receiving 10 or fewer payments for a given HCPCS code were excluded from the dataset to maintain privacy. The National Downloadable File was utilized and linked to the Physician and Other Supplier Public Use File via NPI to obtain year of graduation for each physician^14^. Data from the American Community Survey (ACS) from 2013 to 2019 were used to obtain area-level socioeconomic indicators, such as median household income, unemployment rates, poverty rates and education rates^15^. ACS data were joined to the Physician and Other Suppler Public Use File by ZIP code tabulation areas associated with each physician in the dataset. This analysis did not require IRB approval, as it utilizes publicly available CMS data. All research adhered to the tenets of the Declaration of Helsinki. This is a retrospective study using de-identified subject details. Informed consent was not obtained. The datasets analyzed during the current study are available from the CMS website, https://data.cms.gov/provider-summary-by-type-of-service/medicare-physician-other-practitioners/medicare-physician-other-practitioners-by-provider-and-service and https://data.cms.gov/provider-data/dataset/mj5m-pzi6. The ACS data is available from the United States Census Bureau website, https://www.census.gov/programs-surveys/acs/data.html.

### Study Cohort

This analysis included all physicians who provided services to Medicare beneficiaries between 2013 and 2019. Non-physician healthcare workers were excluded based on the “Credentials” field (e.g., nurse practitioners (NP), registered dieticians (RD)) within the Physician and Other Supplier Public Use File. Gender was obtained from the Physician and Other Supplier Public Use File and coded as man or woman by self-report. Physicians with missing gender data were excluded.

### Study Outcomes

Data were aggregated at the physician-level to obtain the total number of services and total Medicare reimbursement per year, both overall and divided into clinical and procedural services. Total number of services was defined as the total number of services provided by the physician. Total Medicare reimbursement was defined as the sum of the average Medicare standardized payment amount. This represents the average amount that Medicare paid after geographic standardization, removing differences in payment rates for individual services across geographic areas^16^. Clinical services were defined as all HCPCS codes between 99201 and 99499, as well as Eye Visit codes (92002, 92004, 92012, 92014). Procedural services were defined as all other HCPCS codes. J and Q codes were excluded from analysis.

### Statistical Analyses

Statistical analyses were performed using RStudio v1.4.1106. Medians and interquartile range (IQR) were calculated for total number of charges and total CMS payment per year and by gender. Median values were compared using Wilcoxon rank sum tests. To evaluate for differences in reimbursement by gender, we used linear regression models adjusted for total number of services, calendar year, years of experience, reimbursement year, ACS data (median household income, unemployment rates, poverty rates and education rates of workplace), and proportion of total Medicare reimbursement that came from procedural services. P values less than 0.05 were considered statistically significant. We conducted a sensitivity analysis by repeating all analyses after excluding the top and bottom 2.5% of earners overall and in each specialty.

## Results

### Cohort characteristics

Among the 3,831,504 physicians included in this analysis from 2013-2019, 2,712,545 (70.8%) were men and 1,118,859 (29.2%) were women. Of the total number of charges, 55.6% were procedural and 43.4% were non-procedural. The specialties with the highest number of physicians were internal medicine (1,208,422), family medicine (950,252), emergency medicine (451,251), cardiology (331,830), and general surgery (325,983). The states with the highest number of physicians from 2013-2019 were California (70,387 physicians, 68.5% men), New York (56,304 physicians, 66.0% men), Texas (49,901 physicians, 70.1% men), Florida (47,436 physicians, 72.8% men), and Pennsylvania (38,786 physicians, 68.3% men).

Overall, men received more in Medicare reimbursements ($58,815 ± $104,772 vs. $32,205 ± $60,556, p<0.001), billed more services (864 ± 1,780 vs. 505 ± 1,007, p<0.001), and had more years of experience (23 ± 18 vs. 17 ± 15, p<0.001) compared to women **(Table 1, Supplementary Table 1, Supplementary Table 2).** The cumulative difference in total Medicare reimbursements between men and women physicians between 2013 and 2019 amongst physicians who were reimbursed for Medicare all seven years was $188,565,640,740; men physicians were collectively and cumulatively reimbursed $239,667,927,499 while women physicians were cumulatively reimbursed $51,102,286,759. On average, women were reimbursed $482,748 or 58% of what the average man was reimbursed ($831,635) across all seven years.

**Table 1.** Median Medicare reimbursement by gender, 2013-2019.

| | | **N** | **Total Medicare reimbursement (USD\$), median** | **Total Medicare reimbursement (USD\$), IQR** | **p-value** |
| --- | --- | --- | --- | --- | --- |
| **All** | **Men** | 2,712,645 | 58,815 | 104,772 | <0.001 |
|  | **Women** | 1,118,859 | 32,205 | 60,556 |  |
| **2013** | **Men** | 384,790 | 59,710 | 107,373 | <0.001 |
|  | **Women** | 144,250 | 30,575 | 59,631 |  |
| **2014** | **Men** | 385,915 | 57,816 | 102,969 | <0.001 |
|  | **Women** | 149,740 | 30,206 | 57,664 |  |
| **2015** | **Men** | 386,642 | 59,886 | 105,632 | <0.001 |
|  | **Women** | 155,304 | 32,537 | 61,212 |  |
| **2016** | **Men** | 387,986 | 59,604 | 105,124 | <0.001 |
|  | **Women** | 160,873 | 32,616 | 60,976 |  |
| **2017** | **Men** | 388,231 | 58,607 | 103,866 | <0.001 |
|  | **Women** | 164,893 | 32,523 | 60,374 |  |
| **2018** | **Men** | 389,458 | 58,386 | 104,643 | <0.001 |
|  | **Women** | 169,779 | 33,147 | 61,637 |  |
| **2019** | **Men** | 389,623 | 57,874 | 103,837 | <0.001 |
|  | **Women** | 174,020 | 33,456 | 61,762 |  |

### Longitudinal change from 2013-2019

From 2013 to 2019, the number of men and women physicians increased (men: 384,790 to 389,623, women: 144,250-174,020). The ratio of men to women physicians also changed from 2.67 in 2013 to 2.23 in 2019. The median Medicare reimbursement for men physicians decreased from 2013 to 2019 from $59,710 to $57,874, while the median Medicare reimbursement for women physicians increased from $30,575 to $33,456 (**Table 1**, **Figure 1**). The median number of services for men physicians decreased from 2013 to 2019 from 910 to 816, while the median number of services for women physicians remained relatively steady during that time (**Supplementary Table 1, Supplementary Figure 1**).

**Figure 1.**
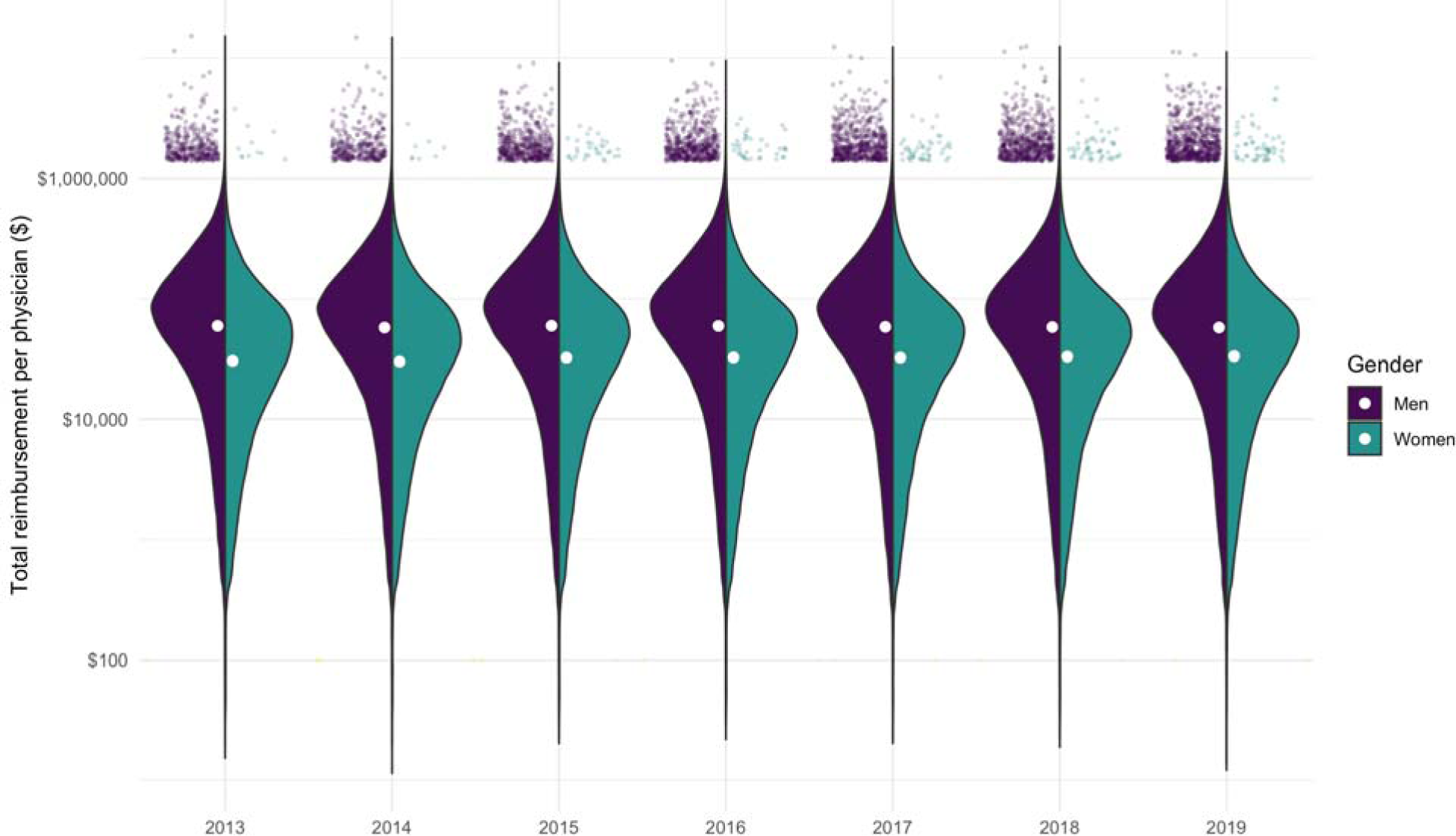
Total reimbursement per physician, 2013-2019. Note: Split violin plot showing the distribution of total reimbursement for each year and gender. The scatter plots at the top display physicians who are reimbursed in the top 0.1%. The white circles represent median reimbursement for that year and gender.

### Differences in specialties

Men physicians were reimbursed more than women physicians in all specialties (**Figure 2**). The specialties with the highest difference in median Medicare reimbursement between men and women were ophthalmology ($99,452), dermatology ($84,844), cardiology ($64,112), nephrology ($62,352), and pulmonary medicine ($47,399). The specialties with the least difference in median Medicare reimbursement between men and women were plastic surgery ($5,806), public health and preventive medicine ($3,639), obstetrics and gynecology ($1,446), anesthesiology ($1,430), and pediatrics ($970).

**Figure 2.**
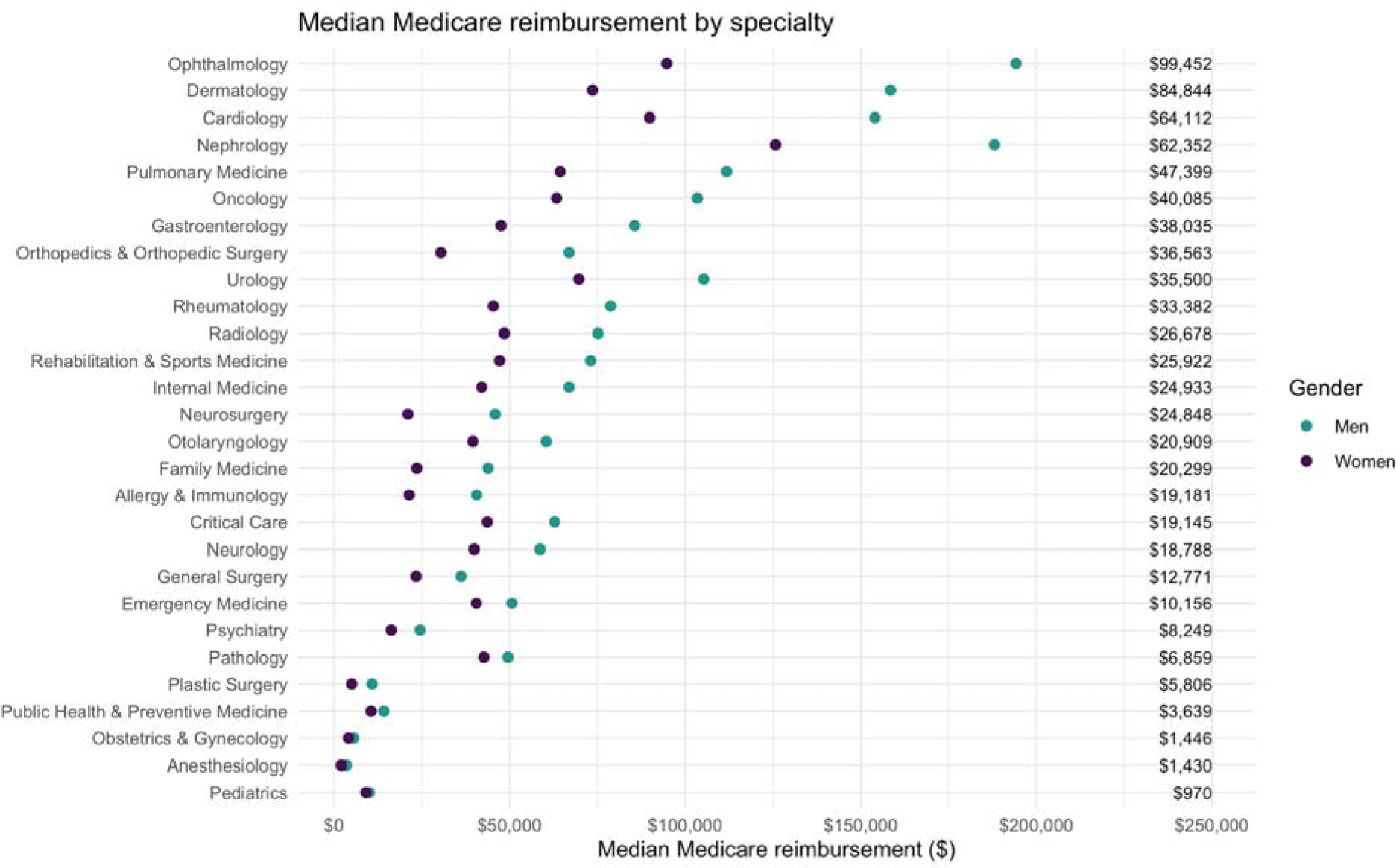
Median Medicare reimbursement by specialty. Note: the column on the right-hand-side shows the difference in median reimbursement between genders per specialty.

In linear regression models controlling for total number of services, calendar year, years of experience, and ACS data (mean household income, unemployment, poverty rate, and education) of place of work, being a man was associated with higher amount of Medicare reimbursement in all specialties, as compared to being women (p<0.01 for all) (**Table 2**). The specialties with the highest estimate difference between men and women physicians after multivariate adjustment were dermatology ($67,160; 95% CI $64,120-$70,200; p<0.01), oncology ($39,010; 95% CI $35,637-$42,384; p<0.01), ophthalmology ($33,147, 95% CI $31,532-$34,762; p<0.01), nephrology ($27,412; 95% CI $24,981-$29,843; p<0.01), and cardiology ($26,337; 95% CI $23,608-$29,607; p<0.01). Several specialties with the highest estimate difference were procedure-heavy, so we additionally adjusted for the proportion of total Medicare reimbursement that came from procedural services for all specialties. After adjusting for procedural services, differences in the gender gap remained significant but were significantly decreased in several procedure-heavy specialties, like dermatology, oncology, ophthalmology, neurosurgery, and orthopedic surgery (**Table 2**).

**Table 2.**
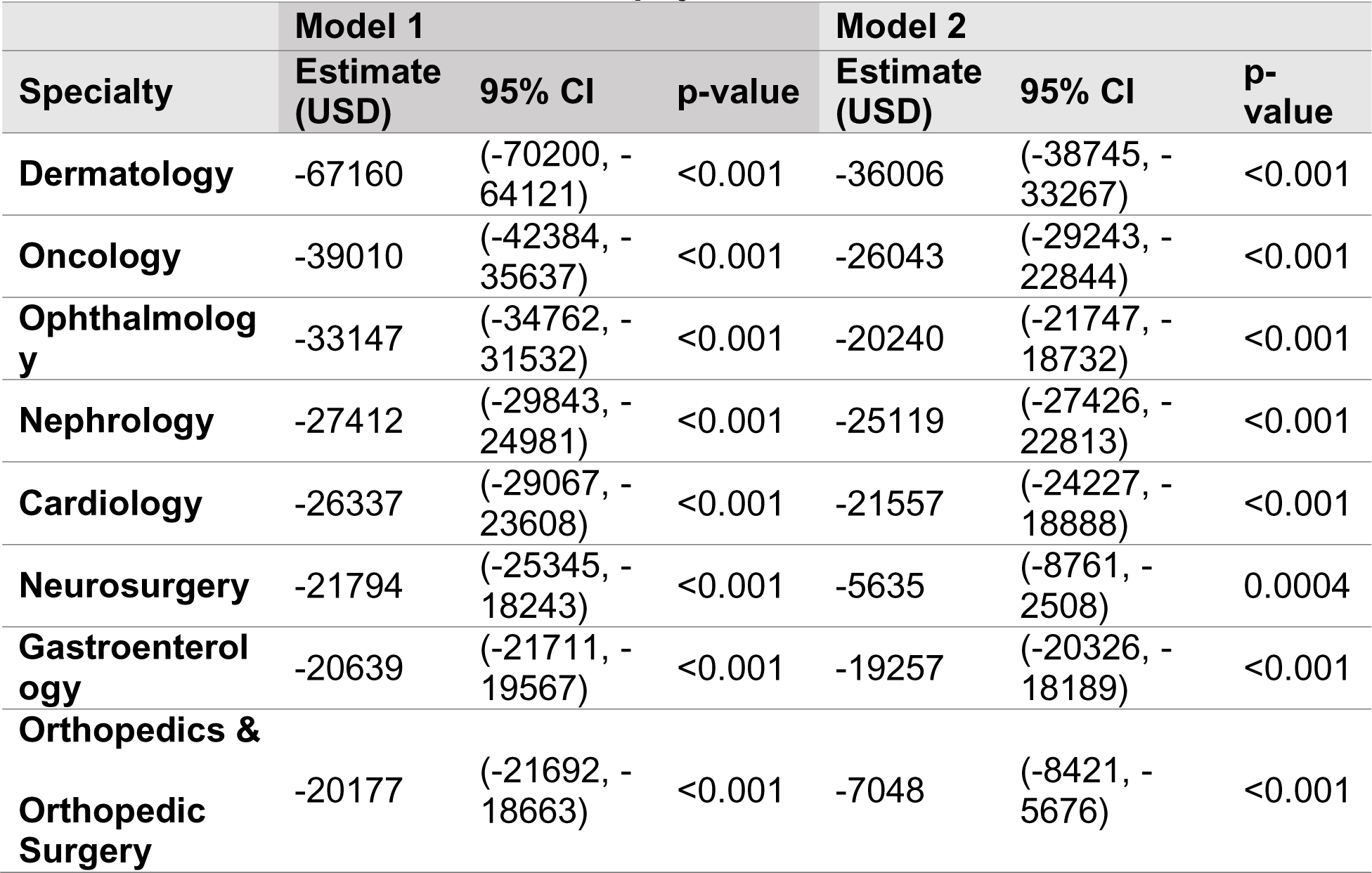
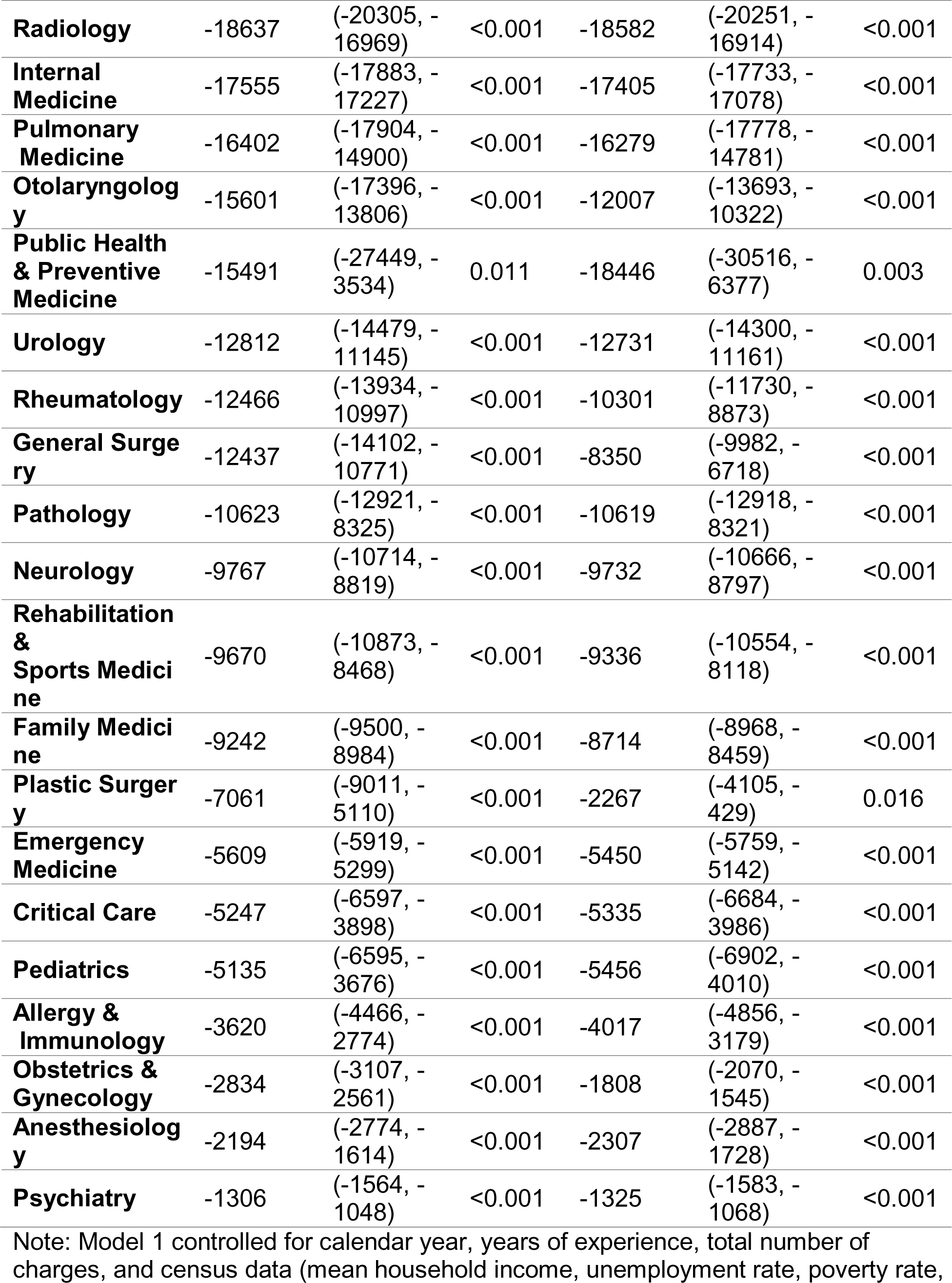
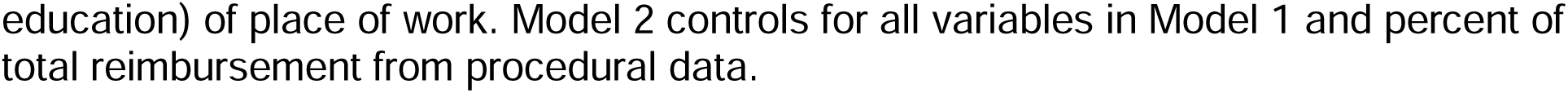
Linear regression for Medicare reimbursement, by specialty *Difference between men and women physicians*.

| **Specialty** | **Model 1** |  |  | **Model 2** |  |  |
| --- | --- | --- | --- | --- | --- | --- |
|  | **Estimate (USD)** | **95% CI** | **p-value** | **Estimate (USD)** | **95% CI** | **p-value** |
| **Dermatology** | -67160 | (-70200, -64121) | <0.001 | -36006 | (-38745, -33267) | <0.001 |
| **Oncology** | -39010 | (-42384, -35637) | <0.001 | -26043 | (-29243, -22844) | <0.001 |
| **Ophthalmology** | -33147 | (-34762, -31532) | <0.001 | -20240 | (-21747, -18732) | <0.001 |
| **Nephrology** | -27412 | (-29843, -24981) | <0.001 | -25119 | (-27426, -22813) | <0.001 |
| **Cardiology** | -26337 | (-29067, -23608) | <0.001 | -21557 | (-24227, -18888) | <0.001 |
| **Neurosurgery** | -21794 | (-25345, -18243) | <0.001 | -5635 | (-8761, -2508) | 0.0004 |
| **Gastroenterology** | -20639 | (-21711, -19567) | <0.001 | -19257 | (-20326, -18189) | <0.001 |
| **Orthopedics & Orthopedic Surgery** | -20177 | (-21692, -18663) | <0.001 | -7048 | (-8421, -5676) | <0.001 |
| **Radiology** | -18637 | (-20305, -16969) | <0.001 | -18582 | (-20251, -16914) | <0.001 |
| **Internal Medicine** | -17555 | (-17883, -17227) | <0.001 | -17405 | (-17733, -17078) | <0.001 |
| **Pulmonary Medicine** | -16402 | (-17904, -14900) | <0.001 | -16279 | (-17778, -14781) | <0.001 |
| **Otolaryngology** | -15601 | (-17396, -13806) | <0.001 | -12007 | (-13693, -10322) | <0.001 |
| **Public Health & Preventive Medicine** | -15491 | (-27449, -3534) | 0.011 | -18446 | (-30516, -6377) | 0.003 |
| **Urology** | -12812 | (-14479, -11145) | <0.001 | -12731 | (-14300, -11161) | <0.001 |
| **Rheumatology** | -12466 | (-13934, -10997) | <0.001 | -10301 | (-11730, -8873) | <0.001 |
| **General Surgery** | -12437 | (-14102, -10771) | <0.001 | -8350 | (-9982, -6718) | <0.001 |
| **Pathology** | -10623 | (-12921, -8325) | <0.001 | -10619 | (-12918, -8321) | <0.001 |
| **Neurology** | -9767 | (-10714, -8819) | <0.001 | -9732 | (-10666, -8797) | <0.001 |
| **Rehabilitation & Sports Medicine** | -9670 | (-10873, -8468) | <0.001 | -9336 | (-10554, -8118) | <0.001 |
| **Family Medicine** | -9242 | (-9500, -8984) | <0.001 | -8714 | (-8968, -8459) | <0.001 |
| **Plastic Surgery** | -7061 | (-9011, -5110) | <0.001 | -2267 | (-4105, -429) | 0.016 |
| **Emergency Medicine** | -5609 | (-5919, -5299) | <0.001 | -5450 | (-5759, -5142) | <0.001 |
| **Critical Care** | -5247 | (-6597, -3898) | <0.001 | -5335 | (-6684, -3986) | <0.001 |
| **Pediatrics** | -5135 | (-6595, -3676) | <0.001 | -5456 | (-6902, -4010) | <0.001 |
| **Allergy & Immunology** | -3620 | (-4466, -2774) | <0.001 | -4017 | (-4856, -3179) | <0.001 |
| **Obstetrics & Gynecology** | -2834 | (-3107, -2561) | <0.001 | -1808 | (-2070, -1545) | <0.001 |
| **Anesthesiology** | -2194 | (-2774, -1614) | <0.001 | -2307 | (-2887, -1728) | <0.001 |
| **Psychiatry** | -1306 | (-1564, -1048) | <0.001 | -1325 | (-1583, -1068) | <0.001 |

Note: Model 1 controlled for calendar year, years of experience, total number of charges, and census data (mean household income, unemployment rate, poverty rate,
education) of place of work. Model 2 controls for all variables in Model 1 and percent of total reimbursement from procedural data.

Similarly, in linear regression models controlling for calendar year, years of experience, and ACS data (mean household income, unemployment, poverty rate, and education) of place of work, being a man was associated with reimbursement for a greater number of services in all specialties except for obstetrics and gynecology (**Supplementary Table 3**). This finding was significant in all specialties except for public health and preventive medicine. The specialties with the highest estimate difference between men and women physicians were allergy and immunology ($1,618; 95% CI $1,489-$1,746; p<0.01), dermatology ($1,456; 95% CI $1,403-$1,510; p<0.01), ophthalmology ($1,065, 95% CI $1,029-$1,102; p<0.01), rheumatology ($942; 95% CI $873-$1,010; p<0.01), and oncology ($820; 95% CI $757-$883; p<0.01). Several specialties with the highest estimate difference were procedure-heavy, so we additionally adjusted for the proportion of total Medicare reimbursement that came from procedural services for all specialties. After adjusting for procedural services, differences in the gender gap remained significant (with the exception of plastic surgery) but were significantly decreased in several procedure-heavy specialties, like allergy and immunology, dermatology, ophthalmology, neurosurgery, rheumatology, and orthopedic surgery (**Supplementary Table 3**).

### Top-earners

We examined the proportion of men and women healthcare physicians who are “top-earners,” defined as those who are reimbursed in the top 1% of physicians within their respective specialties. Overall, men physicians make up 71% of healthcare physicians but comprise of 91% of top-earners. Men physicians were the majority of top-earners in all specialties; the percentage of top earning men physicians (range: 65%-99.5%) surpassed the proportion of men in each specialty (range: 46%-95%), with the exception of public health and preventive medicine **(Supplementary Table 4).** The specialties with the highest percentage of men top-earners were urology (99.5%), orthopedics and orthopedic surgery (99.5%), neurosurgery (99.3%), gastroenterology (97.9%) and allergy and immunology (97.7%). **Figure 1** and **Supplementary Figure 1** also demonstrate that the majority of physicians reimbursed in the top 0.1% or charging in the top 0.1% services are men, though the number of women physicians in this category is increasing annually.

### Geographic variation

Finally, we examined geographic differences in mean Medicare reimbursement and number of charges. **Figure 3** and **Supplementary Figure 4** show the gender difference in total Medicare reimbursement and number of charges in each state. In all states, men were reimbursed and had a greater number of services than women. The states with the highest gender differences in mean reimbursement were Florida ($66,000.63), Delaware ($65,140.93), Alabama ($61,925.30), Mississippi ($61,845.83), and Louisiana ($55,835.57). The states with the lowest gender differences in mean reimbursement were Minnesota ($13,751.97), Wyoming ($14,749.28), Vermont ($16,990.12), Alaska ($17,742.21), and Wisconsin ($21,147.15).

**Figure 3:**
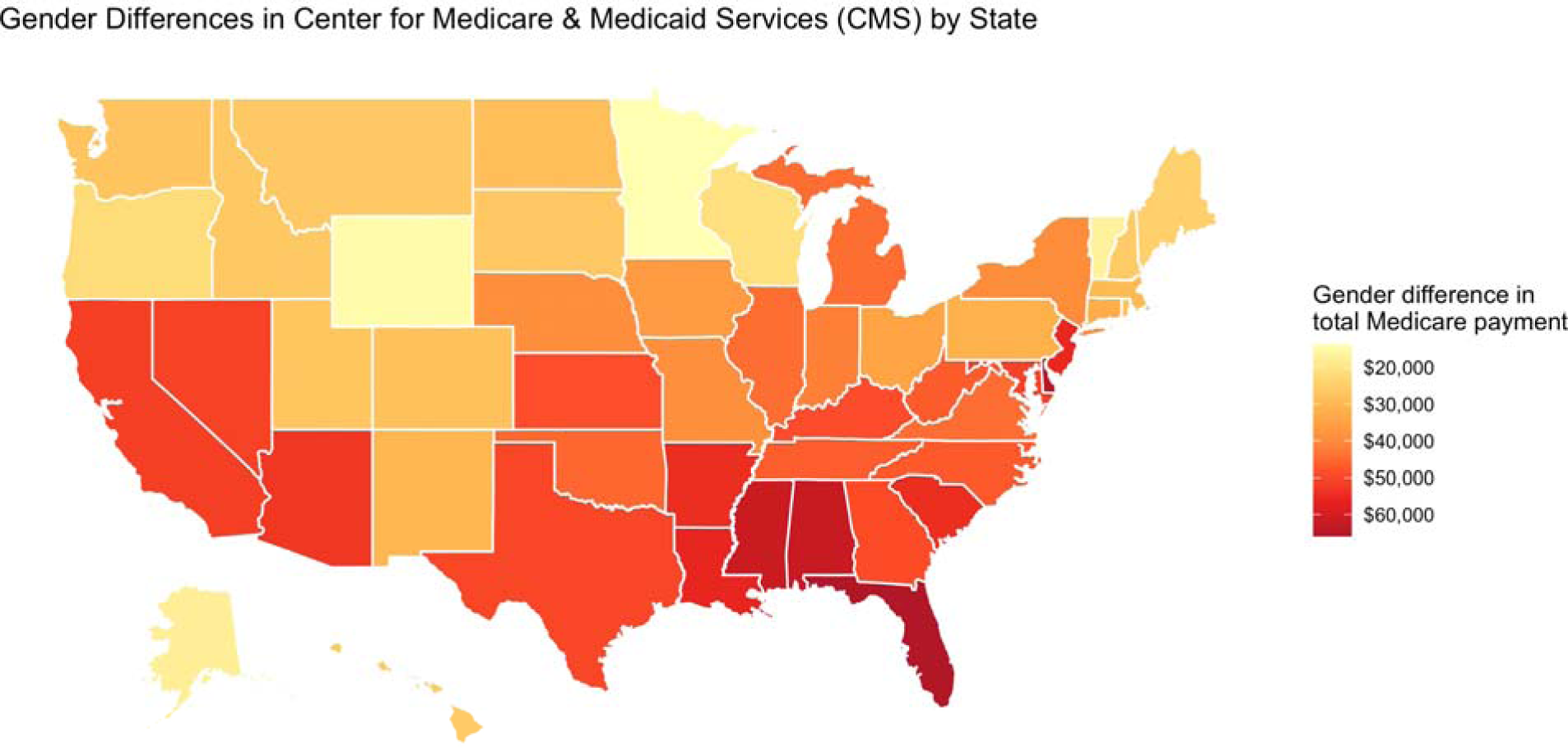
Differences in mean Medicare reimbursement per provider, by state.

## Discussion

Men physicians received higher total annual reimbursements than women and submitted a higher mean number of charges across all medical and surgical specialties. Importantly, after multivariable adjustment for potential confounders including clinical volume (total number of services), differences in total reimbursement remained statistically significant across most specialties. Differences in median reimbursements were highest in procedure-based specialties, including ophthalmology, dermatology, and cardiology. Men accounted for a much higher proportion of the top earners compared to their share of the total Medicare-reimbursed physician population.

Gender based differences in pay have been previously reported in multiple specialties, including in ophthalmology, cardiology and otolaryngology^5,7^. Here we demonstrate this finding spans all medical and surgical specialties, though to a variable extent, and persists over time. Our findings suggest factors that may explain the persistent gender pay gap among physicians. We found that procedure-heavy specialties (e.g., ophthalmology, dermatology, and cardiology) had the largest difference in median Medicare reimbursement. This may be partially explained by the fact that procedure-heavy specialties have higher reimbursements per service provided, thus differences in the number of services provided by men and women physicians may result in a greater disparity. We also found that differences in the volume of procedural services partially explained disparities in total reimbursement; differences in the gender pay gap were significantly reduced in procedure-heavy specialties like ophthalmology, dermatology, neurosurgery and orthopedic surgery after adjusting for the proportion of payments received for procedural services. These results suggest that, within a given specialty, women physicians are less likely to bill for procedural services, and therefore more likely to receive lower reimbursements compared to men. A prior study using Medicare payment data found that men ophthalmologists performed a higher number of cataract surgeries than women ophthalmologists, even after controlling for clinical productivity and years of experience^17^. Among pediatric surgeons in academic medical centers, women surgeons had a lower overall case volume than men, with a lower share of specialist cases^18^.

Additionally, we found that women physicians submitted a lower mean number of charges to Medicare, potentially due to lower clinical volume overall. Indeed, prior studies on differences in Medicare reimbursement by gender in different specialties have similarly found that women physicians received payments for a lower mean number of charges^5,7,8^. These differences in practice patterns may be partially explained by the disproportionate burden of administrative and academic responsibilities placed on women physicians^19^. Many of these additional duties are not reimbursed at the same rate as clinical work and may not lead to promotion. Furthermore, women physicians may be more likely to work fewer hours or part-time in order to fulfill parental or family responsibilities. Prior work has shown that clinical volume among women physicians is disproportionally impacted by time-off taken for childcare responsibilities compared to men physicians^20^. A second hypothesis is that women physicians make personal choices to lower their clinical volume in favor of prioritizing parental responsibilities.

However, without parental leave policies giving equal incentives for both men and women physicians to take time off for childcare, the freedom for women physicians to make these personal choices is limited, making it difficult to test the hypothesis that personal choice is a driver of unequal clinical volume and reimbursement. Equitable parental leave policies may allow both men and women physicians the freedom to take time off for childcare responsibilities, without sacrificing household income and opportunities for promotion. Enacted in some US states, these policies have been shown to improve job continuity for women and increase their employment rates several years after childbirth, with minimal negative impacts on employers^21^. For physicians, such policies may help narrow the gap in clinical volume and reimbursement, and relieve the unequal effects of parental leave on salaries and promotion opportunities.

While men comprised 71% of physicians receiving Medicare reimbursements, they made up 91% of the top 1% of earners. The proportion of top-earners who were men was especially high in procedure-based specialties such as urology, orthopedic surgery, and neurosurgery. These results suggest that women physicians are less likely than men to be among physicians earning the highest reimbursements. Furthermore, differences in reimbursement are associated with a cumulative gap over the seven years of CMS data included in this study, amounting to a difference of $348,887 for each women physician over seven years. If extrapolated to a career of 40 years, this cumulative reimbursement gap reaches $1,993,640 per physician, a figure that is consistent with the $2 million cumulative gap previously reported^4^. While our study has only reported on differences in Medicare reimbursements, a previous study using data on physician salary at US public medical schools have also found a pay gap that persists after adjusting for age, experience, specialty, faculty rank, and measures of research productivity and clinical revenue^1^.

We demonstrate geographic variation in the Medicare reimbursement gap, with states such as Florida, Alabama and Mississippi having the greatest differences in total reimbursements between men and women physicians, and states such as Vermont, Minnesota and Wyoming having the least differences, despite the use of standardized payments controlling for geographic variation in Medicare payments per service. It is possible that these geographic differences in total reimbursements are driven by variation in clinical volume, with the gap in clinical activity being greater in some states than in others. Factors that influence differences in clinical activity such as personal choice, parental leave policies, differences in the proportion of procedural services, and unequal referral patterns may vary by geographic region, depending on state, local and hospital policies.

Strengths of this study include the use of CMS payment data across several years, the inclusion of physicians of all specialties and adjustment for variables that may drive differences in reimbursement including clinical activity, physician characteristics, proportion of procedural services, and area-level socioeconomic factors in practice zip-codes. Total reimbursements were measured using standardized Medicare payments to account for geographic differences in payments per service. The findings of this study should also be interpreted in light of some limitations. Unaccounted for in our analyses were sources of physician income other than Medicare reimbursements, differences in per capita, salary-based and other payment models, as well as differences in practice setting (private, community or academic). Furthermore, variables that may influence clinical activity and total reimbursement such as the implementation of equitable paid parental leave policies, compensation for academic and administrative responsibilities, differences in referral patterns and bias in clinic and operating room scheduling have not been measured.

Our study demonstrates that women physicians billed a fewer number of charges to Medicare and received lower annual reimbursements compared to men physicians between 2013 and 2019. Differences in total reimbursement persisted across time and specialty, and after controlling for physician characteristics, practice patterns and geographic location. This gap was the most pronounced among procedure heavy specialties with the majority of top earning physicians being men. While several drivers of inequitable reimbursements have been hypothesized, including inadequate parental leave policies and the disproportionate distribution of academic and administrative responsibilities, data remains limited. Further work is needed to elucidate the root causes of these disparities that may inform policy changes that promote payment equality among physicians.

## Data Availability

The CMS datasets analyzed during the current study are available from the CMS website, https://data.cms.gov/provider-summary-by-type-of-service/medicare-physician-other-practitioners/medicare-physician-other-practitioners-by-provider-and-service and https://data.cms.gov/provider-data/dataset/mj5m-pzi6. The ACS data is available from the United States Census Bureau website, https://www.census.gov/programs-surveys/acs/data.html.

## Declarations

### Ethics approval and consent to participate

This analysis did not require IRB approval, as it utilizes publicly available CMS data. All research adhered to the tenets of the Declaration of Helsinki. This is a retrospective study using de-identified subject details. Informed consent was not obtained.

### Consent for publication

This is a retrospective study using de-identified subject details. Consent for publication is not required.

## Acknowledgments

None

## References

1. Jena AB, Olenski AR, Blumenthal DM. Sex differences in physician salary in US public medical schools. JAMA Intern Med. 2016;2016(9):1294–1304.

2. Ganguli I, Sheridan B, Gray J, Chernew M, Rosenthal MB, Neprash H. Physician work hours and the gender pay gap—evidence from primary care. N Engl J Med. 2020;383(14):1349–1357.

3. Maher MA, Hayes SN, Shanafelt TD, Sloan JA, Erie JC. Gender differences in physician service provision using Medicare claims data. Mayo Clin Proc. 2017;92(6):870–880.

4. Whaley CM, Koo T, Arora VM, Ganguli I, Gross N, Jena AB. Female physicians earn an estimated $2 million less than male physicians over a simulated 40-year career. Health Affairs. 2021;40(12).

5. Raber I, Al Rifai M, McCarthy CP, et al. Gender differences in Medicare payments among cardiologists. JAMA Cardiol. 2021:e213385.

6. Dubinskaya A, Jackson FI, Labrias PR, Riley B, Shepherd JP. Disparity in Medicare payments by gender and training track in female pelvic medicine and reconstructive surgery. Am J Obstet Gynecol. 2021;225(5):566.e561-566.e565.

7. Reddy AK, Bounds GW, Bakri SJ, et al. Differences in clinical activity and medicare payments for female vs male ophthalmologists.. JAMA Ophthalmol. 2017;135(3):205–213.

8. Valle L, Weng J, Jagsi R, et al. Assessment of differences in clinical activity and Medicare payments among female and male radiation oncologists. JAMA Netw Open. 2019;2(3):e190932.

9. Miller AL, Rathi VK, Burks CA, DeVore EK, Bergmark RW, Gray ST. Assessment of gender differences in clinical productivity and Medicare payments among otolaryngologists in 2017. JAMA Otolaryngol Head Neck Surg. 2020;146(9):1–10.

10. Cohen M, Kiran T. Closing the gender pay gap in Canadian medicine. CMAJ. 2020;192(35):E1011–E1017.

11. Appleby J. Gender pay gap in England’s NHS: little progress since last year. BMJ. 2019;365:I2089.

12. Rad EH, Ehsani-Chimeh E, Gharebehlagh N, Kokabisaghi F, Rezaei S, Yaghoubi M. Higher income for male physicians: findings about salary differences between male and female Iranian physicians. Balkan Med J. 2019;36(3):162–168.

13. Centers for Medicare & Medicaid Services. Medicare provider utilization and payment data: physician and other supplier. Accessed October 12, 2021. https://www.cms.gov/research-statistics-data-and-systems/statistics-trends-and-reports/medicare-provider-charge-data/physician-and-other-supplier.

14. Centers for Medicare & Medicaid Services. National Downloadable File. Accessed October 12, 2021.. https://data.cms.gov/provider-data/dataset/mj5m-pzi6.

15. US Census Bureau. 2009-2019 American Community Survey 5-Year Data. Accessed October 12, 2021.. https://www.census.gov/data/developers/data-sets/acs-5year.html.

16. Centers for Medicare & Medicaid Services. Data Dictionary: Medicare Physician & Other Practitioners - by Provider and Service. Accessed October 12, 2021. https://data.cms.gov/resources/medicare-physician-other-practitioners-by-provider-and-service-data-dictionary.

17. Feng PW, Ahluwalia A, Adelman RA, Chow JH. Gender differences in surgical volume among cataract surgeons. Ophthalmology. 2021;128(5):795–796.

18. Zhang B, Westfal ML, Griggs CL, Hung YC, Chang DC, Kelleher CM. Practice patterns and work environments that influence gender inequality among academic surgeons. Am J Surg. 2019;220(1):69–75.

19. Zhuge Y, Kaufman J, Simeone DM, Chen H, Vleazquez OC. Is there still a glass ceiling for women in academic surgery? Ann Surg. 2011;253(4):637–643.

20. Ly DP, Seabury SA, Jena AB. Hours worked among US dual physician couples with children. JAMA Intern Med. 2015;177(10):1524–1525.

21. Rossin-Slater M, J S. The economic imperative of enacting paid family leave across the United States. https://equitablegrowth.org/the-economic-imperative-of-enacting-paid-family-leave-across-the-united-states/. Accessed May 8th, 2022.

