## Supplement for "Gender based disparities in Medicare physician reimbursement persist across years and specialty"

**Supplementary material:**

**Supplementary Table 1. Median number of services by gender, 2013-2019**

|  |  | N | Total number of services, median | Total number of services, IQR | p-value |
| --- | --- | --- | --- | --- | --- |
| All | **Men** | 2,712,645 | 864 | 1,780 | <0.001 |
|  | **Women** | 1,118,859 | 505 | 1,007 |  |
| 2013 | **Men** | 384,790 | 910 | 1,837 | <0.001 |
|  | **Women** | 144,250 | 505 | 1,011 |  |
| 2014 | **Men** | 385,915 | 882 | 1,772 | <0.001 |
|  | **Women** | 149,740 | 498 | 983 |  |
| 2015 | **Men** | 386,642 | 880 | 1,776 | <0.001 |
|  | **Women** | 155,304 | 510 | 1,012 |  |
| 2016 | **Men** | 387,986 | 873 | 1,775 | <0.001 |
|  | **Women** | 160,873 | 512 | 1,007 |  |
| 2017 | **Men** | 388,231 | 853 | 1,750 | <0.001 |
|  | **Women** | 164,893 | 506 | 997 |  |
| 2018 | **Men** | 389,458 | 837 | 1,792 | <0.001 |
|  | **Women** | 169,779 | 504 | 1,019 |  |
| 2019 | **Men** | 389,623 | 816 | 1,757 | <0.001 |
|  | **Women** | 174,020 | 498 | 1,018 |  |

**Supplementary Table 2. Median years of experience by gender, 2013-2019**

|  |  | N | Years of experience, median | Years of experience, IQR | p-value |
| --- | --- | --- | --- | --- | --- |
| All | **Men** | 2,712,645 | 23 | 18 | <0.001 |
|  | **Women** | 1,118,859 | 17 | 15 |  |
| 2013 | **Men** | 384,790 | 21 | 16 | <0.001 |
|  | **Women** | 144,250 | 16 | 13 |  |
| 2014 | **Men** | 385,915 | 22 | 17 | <0.001 |
|  | **Women** | 149,740 | 16 | 14 |  |
| 2015 | **Men** | 386,642 | 22 | 17 | <0.001 |
|  | **Women** | 155,304 | 16 | 14 |  |
| 2016 | **Men** | 387,986 | 23 | 18 | <0.001 |
|  | **Women** | 160,873 | 17 | 14 |  |
| 2017 | **Men** | 388,231 | 23 | 18 | <0.001 |
|  | **Women** | 164,893 | 17 | 15 |  |
| 2018 | **Men** | 389,458 | 23 | 19 | <0.001 |
|  | **Women** | 169,779 | 17 | 15 |  |
| 2019 | **Men** | 389,623 | 24 | 20 | <0.001 |
|  | **Women** | 174,020 | 17 | 15 |  |

**Supplementary Table 3. Linear regression for number of charges, by specialty**

***Difference between men and women physicians***

|  | Model 1 | | | Model 2 | | |
| --- | --- | --- | --- | --- | --- | --- |
| Specialty | **Estimate** | **95% CI** | **p-value** | **Estimate** | **95% CI** | **p-value** |
| Allergy &  Immunology | -1618 | (-1746, -1490) | <0.001 | -1052 | (-1158, -946) | <0.001 |
| Dermatology | -1457 | (-1511, -1403) | <0.001 | -989 | (-1042, -937) | <0.001 |
| Ophthalmology | -1066 | (-1102, -1029) | <0.001 | -705 | (-740, -671) | <0.001 |
| Rheumatology | -942 | (-1010, -873) | <0.001 | -434 | (-487, -382) | <0.001 |
| Oncology | -820 | (-884, -757) | <0.001 | -631 | (-693, -569) | <0.001 |
| Radiology | -757 | (-793, -720) | <0.001 | -767 | (-803, -730) | <0.001 |
| Cardiology | -725 | (-763, -687) | <0.001 | -654 | (-691, -616) | <0.001 |
| Nephrology | -585 | (-620, -550) | <0.001 | -572 | (-607, -537) | <0.001 |
| Pulmonary  Medicine | -449 | (-483, -416) | <0.001 | -442 | (-474, -409) | <0.001 |
| Internal  Medicine | -407 | (-415, -398) | <0.001 | -389 | (-397, -381) | <0.001 |
| Urology | -400 | (-467, -333) | <0.001 | -377 | (-441, -312) | <0.001 |
| Orthopedics &  Orthopedic  Surgery | -394 | (-418, -369) | <0.001 | -205 | (-229, -182) | <0.001 |
| Family Medicine | -355 | (-363, -347) | <0.001 | -359 | (-367, -352) | <0.001 |
| Rehabilitation & Sports Medicine | -330 | (-359, -301) | <0.001 | -270 | (-300, -241) | <0.001 |
| Otolaryngology | -299 | (-343, -254) | <0.001 | -191 | (-232, -149) | <0.001 |
| Pathology | -289 | (-324, -254) | <0.001 | -290 | (-325, -255) | <0.001 |
| Gastroenterology | -288 | (-302, -274) | <0.001 | -287 | (-301, -273) | <0.001 |
| Neurology | -235 | (-250, -220) | <0.001 | -226 | (-241, -211) | <0.001 |
| Critical Care | -203 | (-228, -177) | <0.001 | -185 | (-210, -160) | <0.001 |
| Psychiatry | -173 | (-182, -165) | <0.001 | -174 | (-182, -165) | <0.001 |
| Neurosurgery | -153 | (-179, -127) | <0.001 | -74 | (-99, -49) | <0.001 |
| Anesthesiology | -127 | (-139, -116) | <0.001 | -138 | (-148, -128) | <0.001 |
| Emergency  Medicine | -112 | (-116, -107) | <0.001 | -112 | (-117, -108) | <0.001 |
| General Surgery | -104 | (-112, -96) | <0.001 | -82 | (-90, -74) | <0.001 |
| Pediatrics | -80 | (-121, -39) | 0.0001 | -67 | (-107, -26) | 0.001 |
| Public Health & Preventive  Medicine | -77 | (-207, 53) | 0.247 | -165 | (-291, -40) | 0.01 |
| Plastic Surgery | -57 | (-76, -38) | <0.001 | -10 | (-28, 8) | 0.2662112 |
| Obstetrics &  Gynecology | 15 | (12, 19) | <0.001 | 26 | (23, 30) | <0.001 |

Note: Model 1 controlled for year, years of experience, total number of charges, and census data (mean household income, unemployment rate, poverty rate, education) of place of work. Model 2 controls for all variables in Model 1 and percent of total reimbursement from procedural services.

**Supplementary Table 4. Top earners, by specialty**

| **Specialty** | **Top earners who are men (%)** | **Proportion of men in specialty (%)** | | **p value** |
| --- | --- | --- | --- | --- |
| **Urology** | 99.5% | | 92.0% | <0.001 |
| **Orthopedics & Orthopedic Surgery** | 99.5% | | 94.6% | <0.001 |
| **Neurosurgery** | 99.3% | | 93.1% | <0.001 |
| **Gastroenterology** | 97.9% | | 83.8% | <0.001 |
| **Allergy & Immunology** | 97.7% | | 66.4% | <0.001 |
| **Cardiology** | 97.5% | | 88.0% | <0.001 |
| **Ophthalmology** | 96.7% | | 77.5% | <0.001 |
| **General Surgery** | 96.2% | | 82.8% | <0.001 |
| **Pulmonary Medicine** | 95.9% | | 81.4% | <0.001 |
| **Critical Care** | 95.5% | | 76.7% | <0.001 |
| **Anesthesiology** | 95.3% | | 81.3% | <0.001 |
| **Nephrology** | 94.7% | | 74.1% | <0.001 |
| **Otolaryngology** | 94.2% | | 85.6% | <0.001 |
| **Plastic Surgery** | 92.6% | | 86.1% | 0.003 |
| **Dermatology** | 92.5% | | 53.5% | <0.001 |
| **Emergency Medicine** | 91.4% | | 73.0% | <0.001 |
| **Internal Medicine** | 91.0% | | 63.3% | <0.001 |
| **Oncology** | 90.9% | | 70.1% | <0.001 |
| **Psychiatry** | 90.8% | | 64.2% | <0.001 |
| **Neurology** | 89.6% | | 69.8% | <0.001 |
| **Family Medicine** | 89.6% | | 61.6% | <0.001 |
| **Radiology** | 88.4% | | 77.9% | <0.001 |
| **Rheumatology** | 86.5% | | 57.8% | <0.001 |
| **Rehabilitation & Sports Medicine** | 84.9% | | 68.8% | <0.001 |
| **Pediatrics** | 79.8% | | 56.5% | <0.001 |
| **Pathology** | 70.2% | | 60.3% | <0.001 |
| **Obstetrics & Gynecology** | 65.2% | | 46.1% | <0.001 |
| **Public Health & Preventive Medicine** | 65.0% | | 69.1% | 0.880 |

**Supplementary Figure 1. Total number of services per physician, 2013-2019**

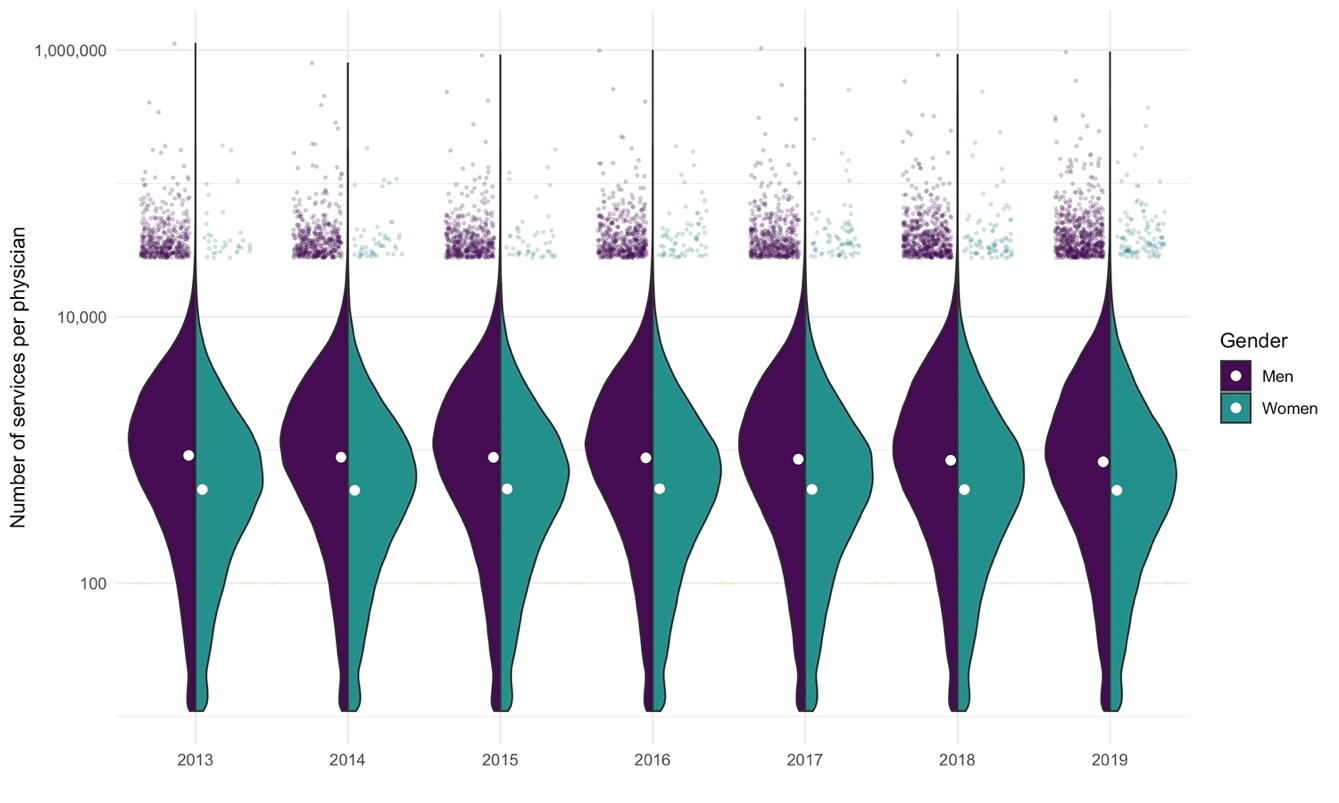

Note: Split violin plot showing the distribution of total reimbursement for each year and gender. The scatter plots at the top display physicians who are reimbursed in the top 0.1%. The white circles represent median reimbursement for that year and gender.

**Supplementary Figure 2. Median Medicare reimbursement by specialty (procedural)**

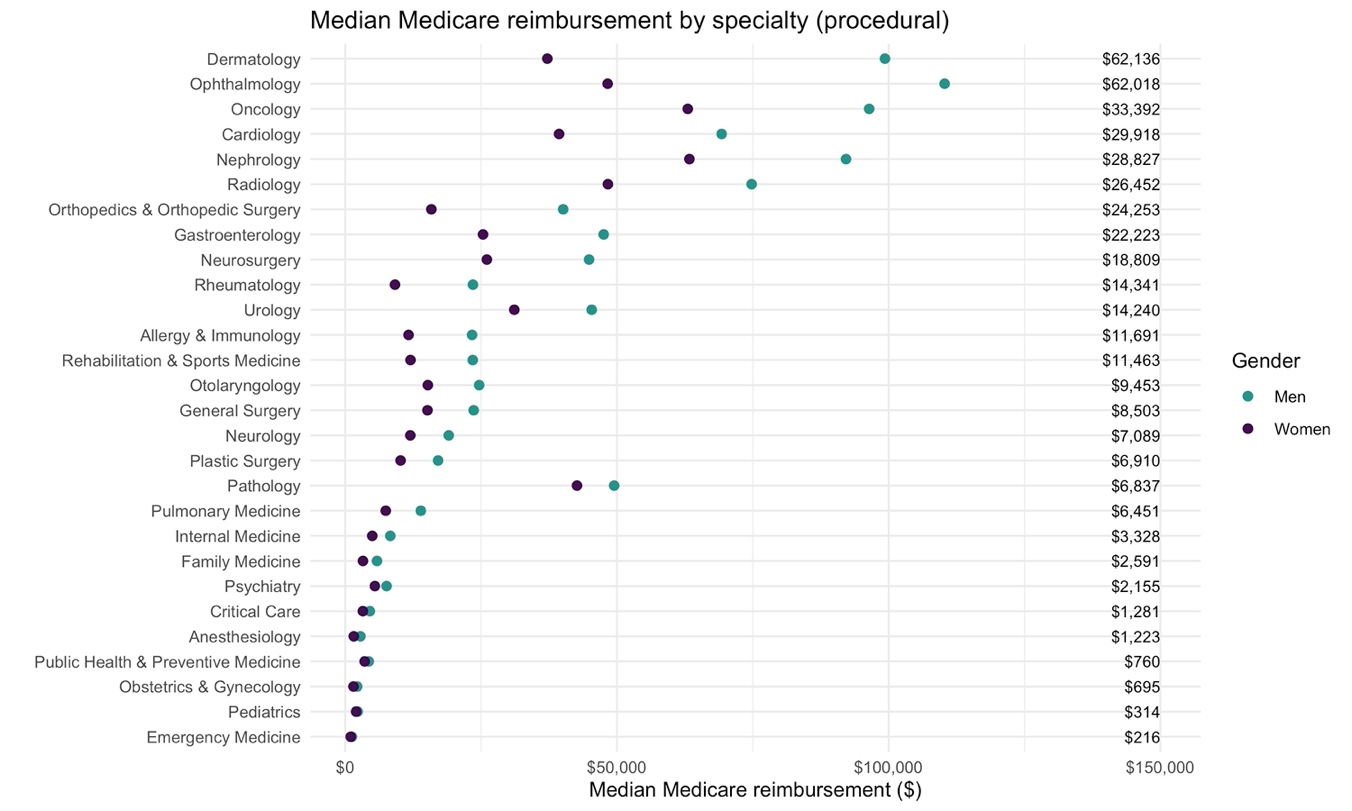

Note: column on the right-hand-side shows difference between median reimbursement between genders per specialty.

**Supplementary Figure 3. Median Medicare reimbursement by specialty (clinical)**

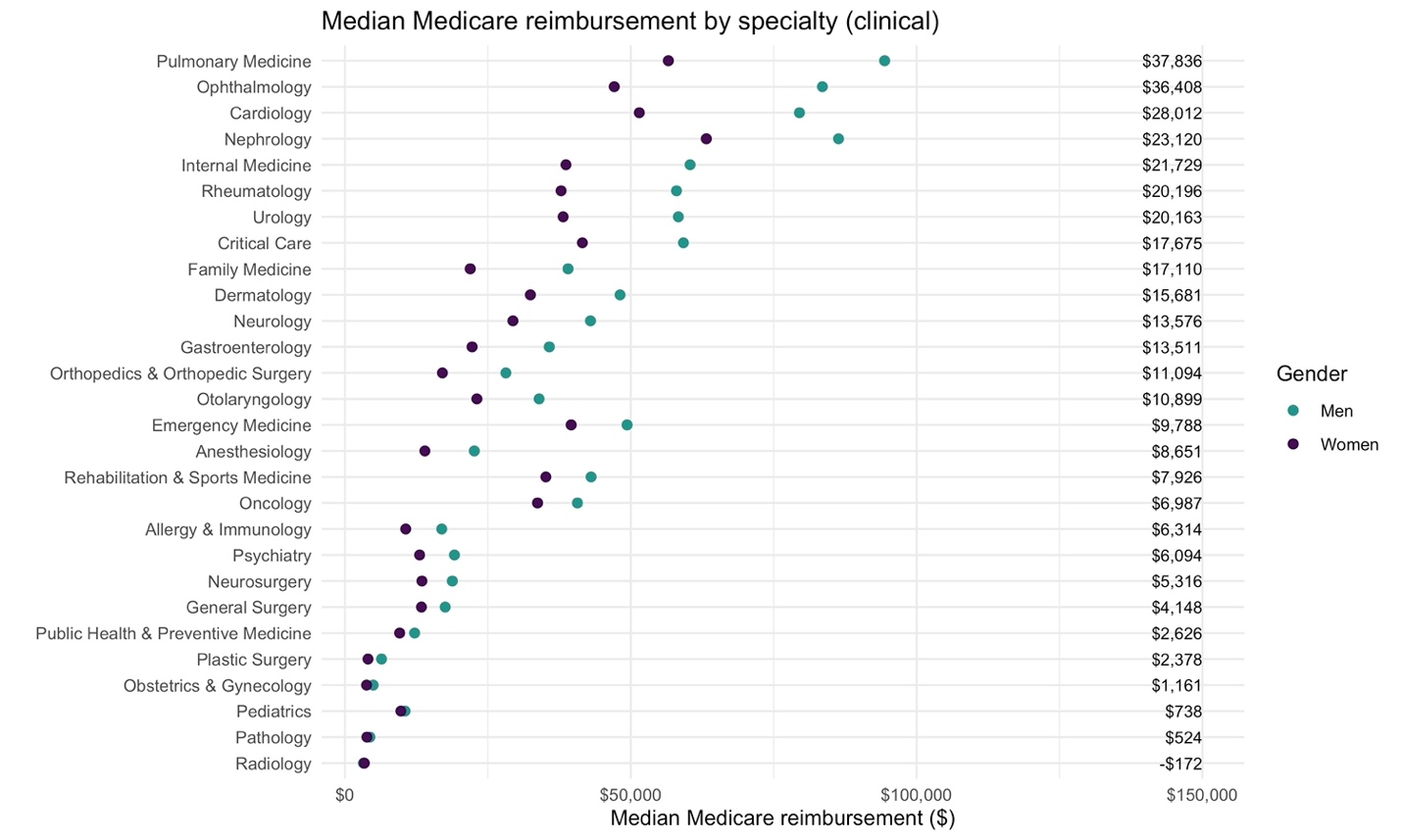

Note: column on the right-hand-side shows difference between median reimbursement between genders per specialty.

**Supplementary Figure 4: Differences in number of Medicare charges per provider, by state**

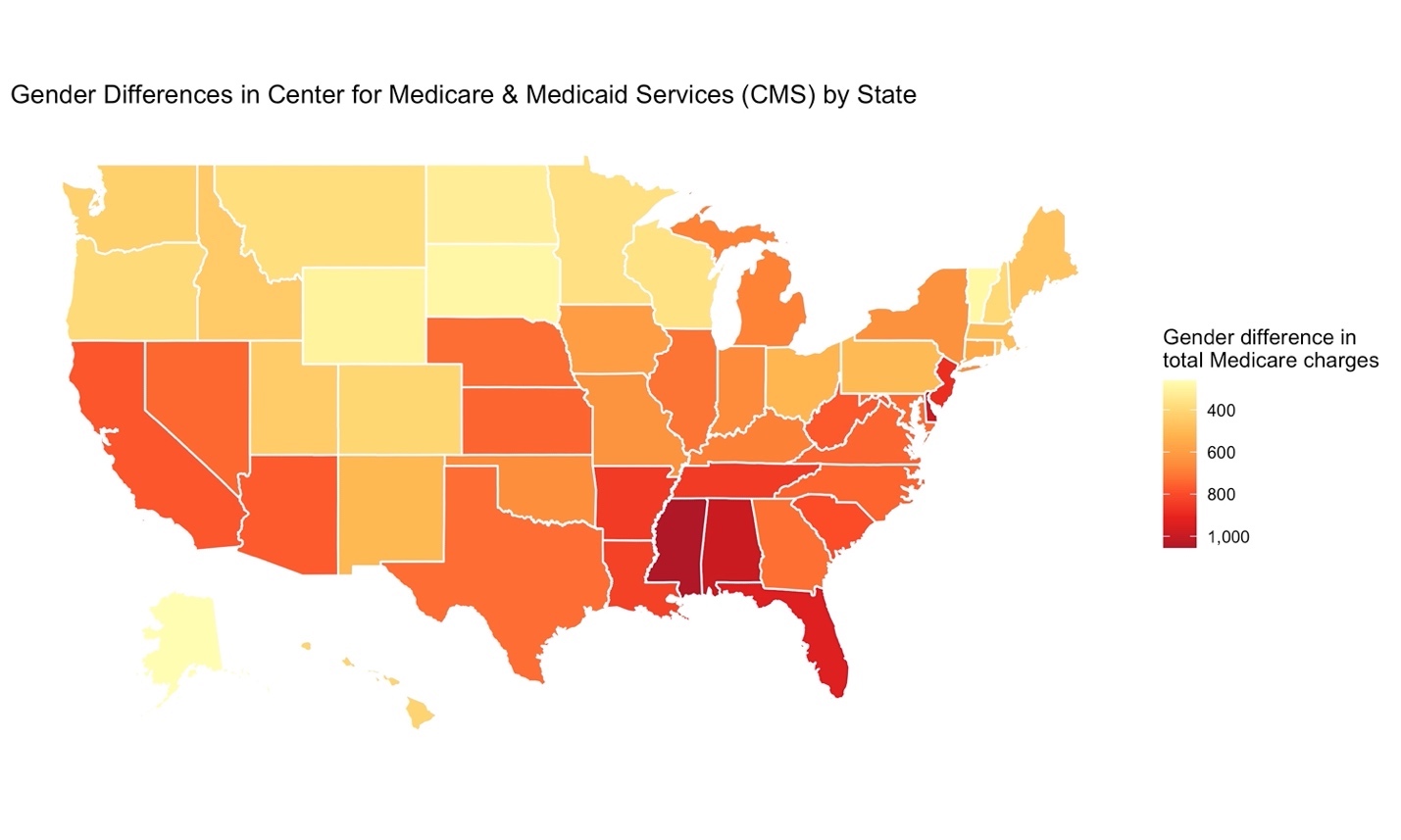
